# Transformation of the Tanzania Medical Store Department through Global Fund support: lessons learned from the field

**DOI:** 10.1101/2020.04.14.20065813

**Authors:** Patrick Githendu, Linden Morrison, Rosemary Silaa, Sai Pothapregada, Sarah Asiimwe, Rafiu Idris, Tatjana Peterson, Emma Davidson, Abaleng Lesego, Neema Mwale, Sako Mwakalobo, Laurean Bwanakunu, Tom Achoki

## Abstract

**Background:** The Tanzania government sought support from The Global Fund to Fight AIDs, Tuberculosis and Malaria (Global Fund) to reform its Medical Stores Department (MSD), with the aim of improving performance. Our study aimed to assess the impact of the reforms and document the lessons learned.

**Methods:** We applied quantitative and qualitative research methods to assess the impact of the reforms. The quantitative part entailed a review of operational and financial data covering the period before and after the implementation of the reforms. We applied interrupted time series analysis to determine the change in average availability of essential health commodities at health zones. Qualitative data was collected through 41 key informant interviews. Participants were identified through stakeholder mapping, purposive and snowballing sampling techniques, and responses were analyzed through thematic content analysis.

**Results:** Availability of essential health commodities increased significantly by 12.6% (95%CI, 9.6-15.6), after the reforms and continued to increase on a monthly basis by 0.2% (95%CI, 0.0-0.3) relative to the preintervention trend. Sales increased by 56.6% while the cost of goods sold increased by 88.6% between 2014/15 and 2017/18. Surplus income increased by 56.4% between 2014/15 and 2017/18, with reductions in rent and fuel expenditure. There was consensus among participants that the reforms, were instrumental in improving performance of MSD.

**Conclusion:** Many positive results were realized through the reforms at MSD. However, despite the progress, there were risks such as the increasing government receivable that could jeopardize the gains. Multi-stakeholder efforts are necessary, to sustain the progress and expand public health.

## Introduction

The World Health Organization (WHO) has clearly elaborated the six core functions of a competent health system. These include; 1) ensuring adequate health financing; 2) development and deployment of human resources for health; 3) effective service delivery; 4) promoting access to essential medicines and other health technologies; 5) collection, analysis and utilization of critical health information; and 6) effective leadership and governance. For impactful delivery of health services, all the six components must work in tandem to produce the desired health outcomes [1, 2]. The Tanzania government has clearly recognized the critical role that access to safe and high-quality medicines and other health technologies play, in ensuring optimal population health outcomes. To this end, MSD was set up as an autonomous department of the Ministry of Health, Community Development, Gender, Elderly and Children (MoHCDGEC). Its overarching mandate is to procure, store and distribute medicines and supplies to all public health facilities in the country [3-8].

Since its founding in 1994, MSD registered steady growth in its business operations, but started to flounder around the year 2006, resulting in frequent stock-outs of essential medicines and other supplies at public health facilities. At this stage, it was evident that the MSD was facing several financial and operational challenges that hindered its effective functioning and delivery on its mandate within the healthcare system [9,10]. Consequently, the Tanzania government sought support of the Global Fund, which commissioned a detailed assessment to comprehensively understand the challenges facing MSD and their underlying causes. This formed the basis of the strategic reforms that were initiated at MSD with the support of the Global Fund, aimed at transforming the organization into an effectively functioning autonomous department within the MoHCDGEC [9-13].

There is evidence from the literature that procurement and supply chain systems for medicines and health commodities in many low- and middle-income countries are faced with end to end challenges [14,15]. These include, problems related to forecasting and quantification; storage, inventory management and distribution; quality assurance, information management and reporting, among others. For instance, Cameron A., et al [16], reports that average availability of generic medicines in the public sector across WHO regions was low, ranging from 29·4% to 54·4%. Purchasing and distribution inefficiencies and price markups leading to significant unaffordability were cited as some of the key bottlenecks. Bigdeli M. et al [17] and Ewen M. et al [18], further support this observation, highlighting the fact that procurement and supply chain challenges contribute significantly to low access to essential medicines and general inefficiency of health systems in many countries. Citing examples from Kenya and Tanzania, Mackintosh M., et al [19], further emphasize that fragmented procurement and distribution systems have negative implications on overall population access to high quality medicines and related health outcomes.

It is based on this understanding that the Global Fund 2017-2022 Strategy clearly acknowledges that building resilient and sustainable health systems, including procurement and supply chain are central to progress towards universal health coverage and acceleration to end the epidemics associated with HIV/AIDS, Tuberculosis and Malaria [20]. To highlight the great importance of this effort, it is estimated that approximately 40% of the Global Fund support going to countries for various disease programs, is used for procurement and supply-chain management of health products [14,15,20]. Therefore, a thorough examination of the lessons drawn from the Tanzania MSD experience, would provide an investment case for various health system stakeholders and donors that are keen to align their strategic funding to implementation objectives, in order to sustainably strengthen health systems. Our article aims to assess the impact of the MSD reforms and document the experiences and lessons learned.

## Methods

This study employed both quantitative and qualitative research methods in assessing the impact of the reforms implemented at the MSD through Global Fund support. The quantitative section focused on trend analysis of priority indicators derived from financial statements and operational reports at MSD. Meanwhile, the qualitative approach entailed key informant interviews that collected and analyzed views from various levels of the healthcare system, in order to give perspective to the observed trends. The overall duration of the study was from July 2019 to September 2019.

## Setting

The Tanzania health system follows a decentralised model, from village dispensaries and community-based health facilities (under the responsibility of local government authorities), to ward, district, and regional level hospitals; with referral and national hospitals at the pinnacle. MSD is responsible for the procurement, storage and distribution of health commodities to an estimated 5,940 health facilities across the country [7,9]. To enhance its efficiency and national coverage, MSD has a network of 8 zonal stores and 2 sales points that service health facilities across the country. Furthermore, within 5 zones, there are 7outlets where members of the community can purchase essential medicines directly [9].

MSD’s procurement function is centralized at the headquarters in Dar es Salaam, which serves as the central warehouse, and as a distribution hub to the respective zones and sales points in the country [9-11]. The warehouse, with an estimated storage capacity of 20,335 square meters was built with support from the U.S. Government and the Global Fund. As a result of increased storage capacity and expanding portfolio of items procured and distributed, MSD increased the items it routinely tracked from 135 to 312 [9, 12,13].

## Intervention

In 2015, the Tanzania government sought assistance from the Global Fund, to undertake a strategic review to understand the main challenges facing MSD that had led to erosion of working capital and subsequently triggered a cycle of operational deficiencies. Subsequently, a set of strategic reforms were successively recommended aimed at improving the overall performance and long-term sustainability of MSD [9-11].

Briefly, the reforms focused on aspects of MSD governance, operational and financial performance. In terms of governance, the Global Fund supported system strengthening by establishing board level oversight capacity and the creation of the Strategic Management Office (SMO), starting from August 2016. The SMO assumed the responsibility of coordination and monitoring various aspects of implementation towards the envisaged strategic reform objectives. More specifically, this entailed training of staff, building technical capacity and development of standard operating procedures and reporting tools, among others. Further, the Global Fund supported the integration of supply-chain training modules into the national curriculum of the health workforce in Tanzania [9,11-13].

In terms of operational performance, Global Fund support was mainly focused on fleet improvements at MSD, which started from January 2017 and continued for the rest of the year. As per the recommendations of a study financed by the Global Fund, a modern fleet of 181 vehicles was procured, to allow for direct delivery of health commodities from zonal stores to respective health facilities [9]. In addition, steady efforts were made towards a comprehensive logistic system re-design, aimed at enhancing efficiencies in the procurement and distribution of medicines and health commodities in the country [9-11]. For instance, at the time of this study, measures were underway to standardize laboratory systems across the country, in order to benefit from the economies of scale in the procurement of equipment and consumables as well as maintenance.

To empower MSD with robust information management systems and capacity to optimize on business processes, an enterprise resource planning (ERP) software, EPICOR, was procured and operationalized through U.S Government assistance. Additional upgrades of EPICOR were made through the Global Fund support in the ensuing periods. Further, in order to ensure financial sustainability of operations, the Global Fund consistently paid an additional 6% of the cost of the medicines and other health commodities it distributes through the MSD channels, to cover the handling and distribution costs. This forms a significant source of revenues for MSD to finance its critical operations [9-11].

### Phase 1: Qualitative research

Phase 1 of the study applied qualitative research methods and collected views of key informants who were knowledgeable and intimately involved in the reform initiative at MSD.

### Sampling and data collection

Participants were selected through non-probability sampling procedures termed purposive sampling and supplemented through snowballing until we reached information saturation [21, 22]. In our case, purposive sampling was a judgement selection based on the participant’s knowledge and involvement in the MSD reform through Global Fund support.

In order to guide our initial judgement in the identification and selection of study participants, we first undertook a comprehensive desk review mapping out the different stakeholders involved in the strategic reforms at MSD. A representative list of 17 organizations, including the public, private, development partners and civil society organizations were drawn to ensure that views of all key stakeholders were represented in our study. Briefly, the public sector organizations included, MoHCDGEC, MSD, President’s Office Regional and Local Government (PORALG), and representatives of health facilities. The private sector organizations included, Deloitte Consulting and PricewaterhouseCoopers, Tanzania (as the Local Funding Agency); while the development partners included, Global Fund, USAID-Global Heath Supply Chain Program, United Nations agencies and WHO.

From the list of organizations identified in the desk review, key informants were identified based on their functions and knowledge of the reform at MSD. In total, we identified 47 participants of which 6 were not able to participate, mainly due to time and logistical constraints. Table 1 provides a breakdown of the different categories of key informants.

**Table 1:** Categories of key informants.

| Sector | Category of participant | Number |
| --- | --- | --- |
| **Public sector** | Policy maker | 4 |
|  | Management | 12 |
|  | Frontline health workers | 10 |
| **Private sector** | Management | 3 |
| **Development partners** | Management | 7 |
| **Civil society** | Management | 5 |
| **Total** |  | **41** |

The interview process used a semi-structured interview guide developed to collect views of the participants. This was first developed in English and then translated into Swahili, which is the national language in Tanzania. The interview guide was comprehensively pretested and necessary modifications and adjustments were made before field deployment. The key informant interview (KII) approach, which involved in-depth interviews was utilized to collect data from the selected participants.

To conduct the interview, arrangements were made to secure a 1-hour appointment with the respective participant, at a suitable venue for a face-to–face discussion. Prior to starting the interview, researchers introduced themselves, explaining the objectives of the study and obtained verbal informed consent to proceed with the interview. Participants were made aware that they could stop participating in the interview at any stage without prejudice. The questions in the interview guide served to sign post the conversation and ensured that respondents covered all aspects of interest in our research.

### Data analysis

Qualitative data obtained from interviews was transcribed and then analyzed applying thematic content analysis. In this approach, data from interview transcripts was grouped into similar concepts. We found this approach to be appropriate for semi-structured expert interviews as it is used for coding text with a predefined coding system, which can then be refined and complemented with new emerging themes.

#### Phase 2: Quantitative research

The quantitative component of the study entailed a detailed analysis of the operational and financial indicators, reported by MSD covering a period from January 2015 to December 2019, with a reform intervention starting on August 2016. Our analysis examined trends of priority indicators, before and after the implementation of the reform, in order to track the resultant changes in MSD operations and financial position.

To determine the impact of the intervention on the availability of essential health commodities, we applied interrupted time series analysis (ITSA), using a user-written STATA command of “ITSA” which uses OLS regression-based models specifically designed for time-series data. Our model of choice was Newey-West, which is suited to handle autocorrelation in addition to possible heteroskedasticity [23].

We set the time point of the intervention at December 2016 in our primary analysis, which was the midpoint of the intervention component aimed at setting up the SMO, and the beginning of the fleet improvements at MSD. Our analysis assumed a 2-month potential time lag partly because according to conversations with MOH experts, this is the time it normally took for projects of this nature to be planned and executed within the health system in Tanzania.

The regression model used in ITSA is represented by the equation [23]:

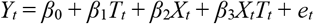

*Where*,

- Y_t_ is the aggregated outcome variable measured at each equally spaced time point t,
- T_t_ is the time since the start of the study,
- X_t_ is a dummy variable representing the intervention (preintervention periods 0, otherwise 1)
- X_t_T_t_ is an interaction term, between X and T.
- e_t_ is the estimate of the error
- *β*_0_ represents the intercept or baseline of the outcome variable.
- *β*_1_ is the slope or trajectory of the outcome variable until the introduction of the intervention.
- *β*_2_ represents the change in the level of the outcome that occurs in the period immediately following the introduction of the intervention (compared with the counterfactual).
- *β*_3_ represents the difference between preintervention and postintervention slopes of the outcome.

To adjust for seasonality and other long-term data trends, we fitted the data on availability of health commodities to a cubic spline over time to cover the full time series. The predicted results from the cubic spline are show on appendix 1. We then analyzed the predicted values using the same ITSA model described above [24]. Additionally, to assess potential different time lags regarding the impact of the reform, we performed sensitivity analyses of the ITSA with various interrupted time periods: from October 2016 to February 2017 (results not shown), besides the primary ITSA analysis, and the results were robust. All p-values were two-sided, and p < .05 was considered statistically significant. Data were analyzed using STATA/MP16.0 (StataCorp LP, College Station, TX).

### Ethical considerations

Relevant permissions and authorization to conduct this research were sought and obtained from MSD and MoHCDGEC. The data collection process ensured that all participants fully understood the objectives of the study and consented verbally to provide the required information.

### No Patient and Public Involvement

This research was done without patient or public involvement.

## Results

### Operational Performance

Average availability of the 312 trace medicines and health commodities at the MSD stores in the preintervention period was 63.5% (95 CI, 62.6-64.1), but increased to 79.9% (95% CI, 78.9 −81.1) in the postintervention period. Figure 1 shows the interrupted time series analysis, with actual data points and predicted results, before and after the intervention, divided by the vertical line.

**Figure 1:**
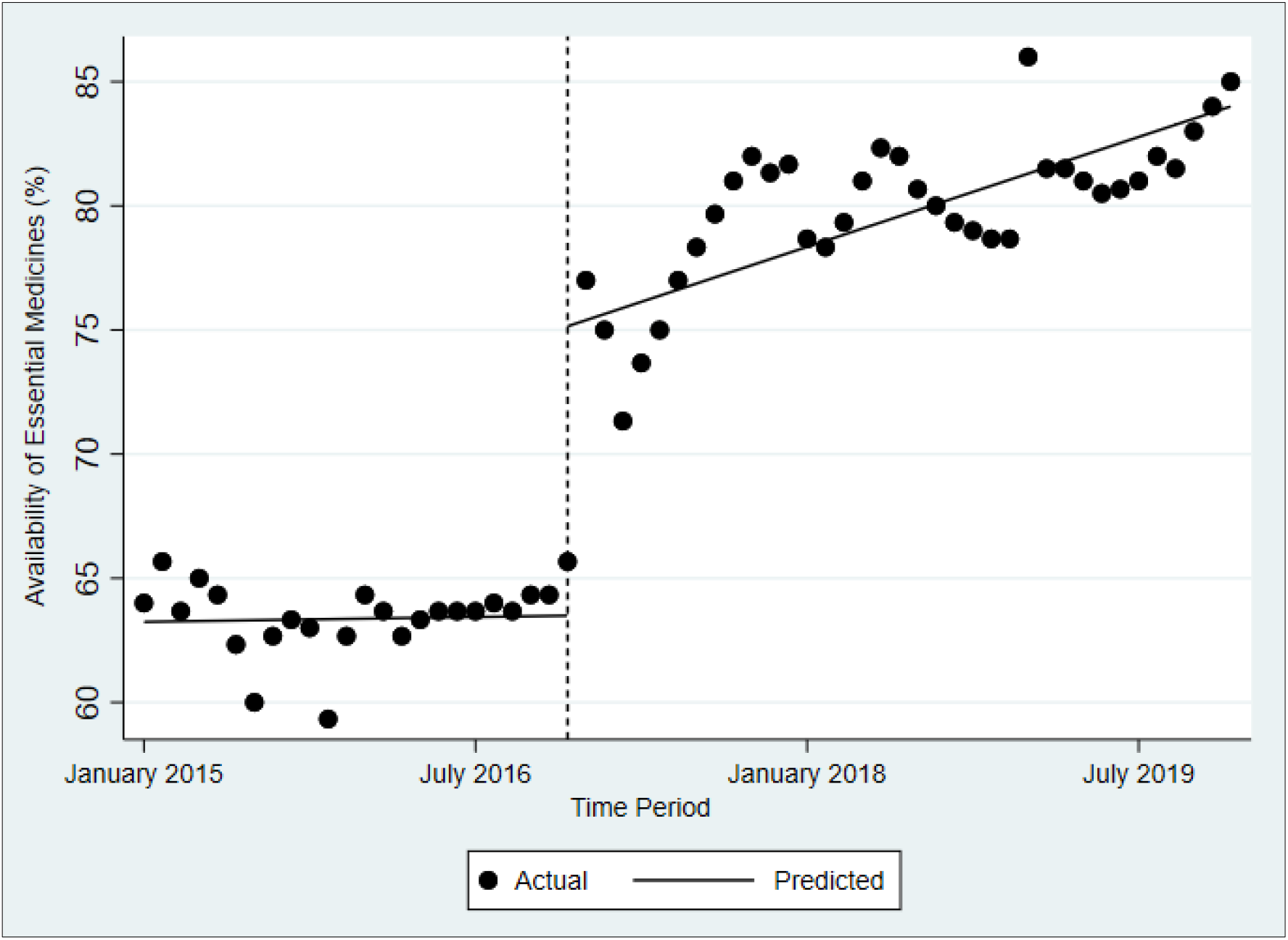
Interrupted time series analysis.

Table 2 shows the regression results using data that is unadjusted for seasonality (A), and data adjusted for seasonality (B). The starting level of availability of health commodities based on unadjusted data was estimated at around 63.1 %; but this appeared to increase every month by about 0.03%, until the beginning of the intervention; although the increase was not significant. In the period immediately after the intervention, availability increased significantly by 12.6% (95%CI, 9.6-15.6), and continued to increase on a monthly basis by 0.2% (95%CI, 0.0-0.3) relative to the pre-intervention trend. Further, the post-intervention trajectory indicated that availability significantly increased by 0.2% (95%CI, 0.1-0.3) each month.

**Table 2:**
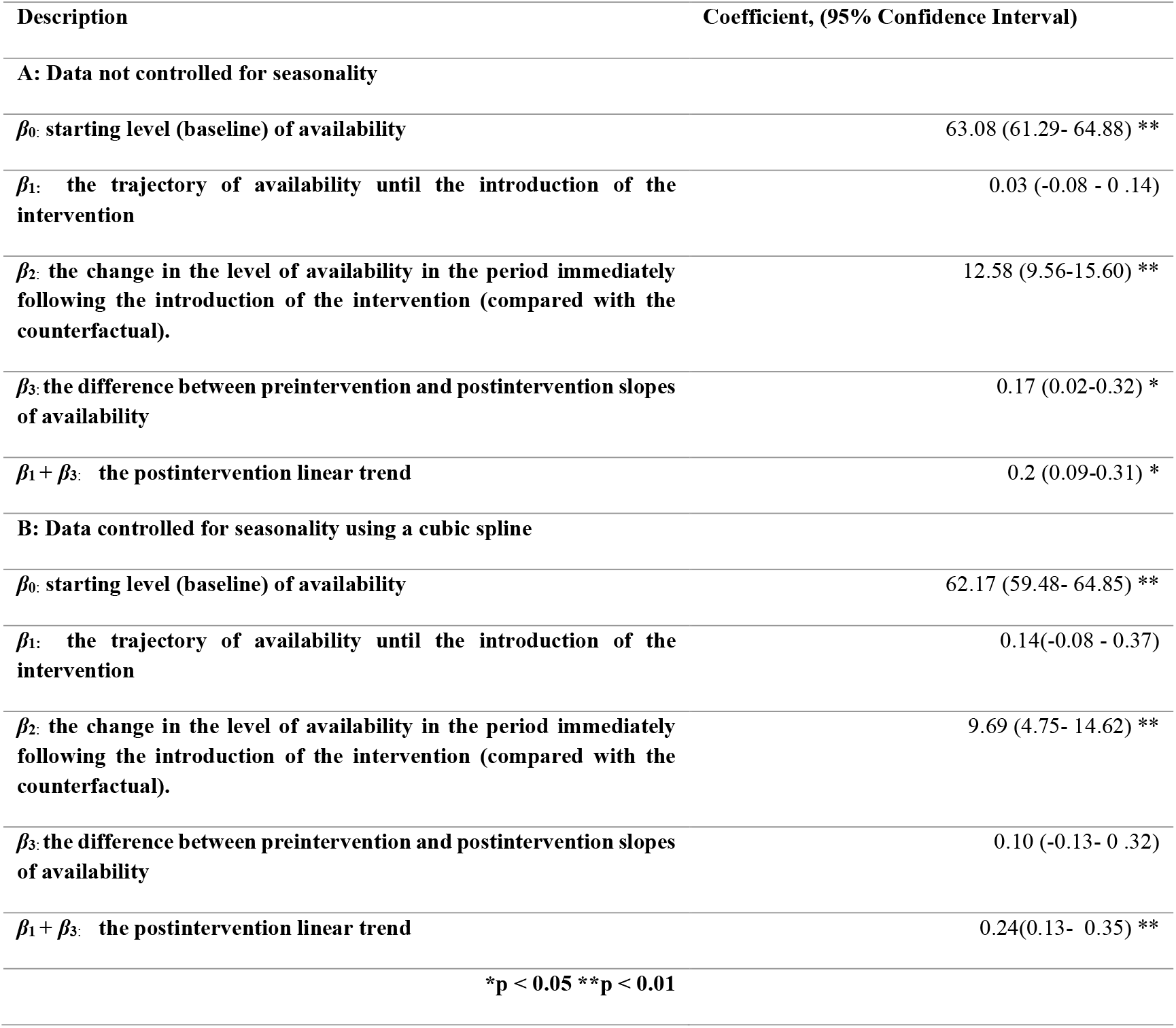
Coefficients estimated from interrupted time series analysis models.

| Description | Coefficient, (95% Confidence Interval) |
| --- | --- |
| **A: Data not controlled for seasonality** |  |
| $\beta_0$: starting level (baseline) of availability | 63.08 (61.29- 64.88) ** |
| $\beta_1$: the trajectory of availability until the introduction of the intervention | 0.03 (-0.08 - 0.14) |
| $\beta_2$: the change in the level of availability in the period immediately following the introduction of the intervention (compared with the counterfactual). | 12.58 (9.56-15.60) ** |
| $\beta_3$: the difference between preintervention and postintervention slopes of availability | 0.17 (0.02-0.32) * |
| $\beta_1 + \beta_3$: the postintervention linear trend | 0.2 (0.09-0.31) * |
| **B: Data controlled for seasonality using a cubic spline** |  |
| $\beta_0$: starting level (baseline) of availability | 62.17 (59.48- 64.85) ** |
| $\beta_1$: the trajectory of availability until the introduction of the intervention | 0.14(-0.08 - 0.37) |
| $\beta_2$: the change in the level of availability in the period immediately following the introduction of the intervention (compared with the counterfactual). | 9.69 (4.75- 14.62) ** |
| $\beta_3$: the difference between preintervention and postintervention slopes of availability | 0.10 (-0.13- 0.32) |
| $\beta_1 + \beta_3$: the postintervention linear trend | 0.24(0.13- 0.35) ** |
| *p < 0.05 **p < 0.01 |  |

Using seasonally adjusted data, the starting point of availability of health commodities was estimated at 62.2%. However, availability appeared to increase significantly by 10.0% (95%CI, 4.8-14.6), in the period immediately after the start of the intervention. Moreover, there was a 0.1% (95% CI, −0.1 – 0.3) monthly increase relative to the pre-intervention period, but this was not significant. The post-intervention trajectory indicated a monthly increase in availability by 0.2% (95%CI, 0.1-0.4).

Table 3 shows the quarterly order fill rates for all medicines and commodities requested at different levels of the health system between 2015 and 2018. On average, the order fill rate from the zones to facilities, dropped from 63.8% to 55.0% between the reporting periods 2015/16 and 2017/18. Meanwhile, the order fill rate from the central medical warehouse to the zones, reported a 26.5% decline over the same reporting period. However, table 3, also shows that the integrated logistics system (ILS) and the related delivery of commodities attained consistently high coverage ranging between 85% and 100%, over the same period.

**Table 3:** Quarterly operational performance.

| Measure | 2015/16 Actual Performance |  |  |  | 2016/17 Actual Performance |  |  |  | 2017/18 Actual Performance |  |  |  |
| --- | --- | --- | --- | --- | --- | --- | --- | --- | --- | --- | --- | --- |
|  | Q1 | Q2 | Q3 | Q4 | Q1 | Q2 | Q3 | Q4 | Q1 | Q2 | Q3 | Q4 |
| **Order fill rate (from zone to facility)** | 57.2% | 63.3% | 62.7% | 73.4% | 69.5% | 63.3% | 63.6% | 61.3% | 57.0% | 53.9% | 54.2% | 56.5% |
| **Order fill rate (from HQ to Zones)** | 98.7% | 95.5% | 52.6% | 46.8% | 32.0% | 31.0% | 42.0% | 32.7% | 40.6% | 48.1% | 54.4% | 72.8% |
| **ILS Delivery coverage** | 95.3% | 91.5% | 85.7% | 92.3% | 92.4% | 89.4% | 97.2% | 95.8% | 93.7% | 97.1% | 100% | 100% |

### Financial Performance

Figure 2 shows that between the financial reporting periods 2014/2015 and 2018/2019, sales increased by 41.2% while the cost of goods sold (COGS) increased by 60.9%. The gross profit margins remained stable over the same period, showing a slight improvement of approximately 3.2% between 2014/2015 and 2018/19. However, the peak sales were in the period 2017/2018, which registered a 56.6% increase from the 2014/2015 baseline. There was a commensurate increase in COGS of 88.6% over the same period, with gross margins remaining relatively stable.

**Figure 2:**
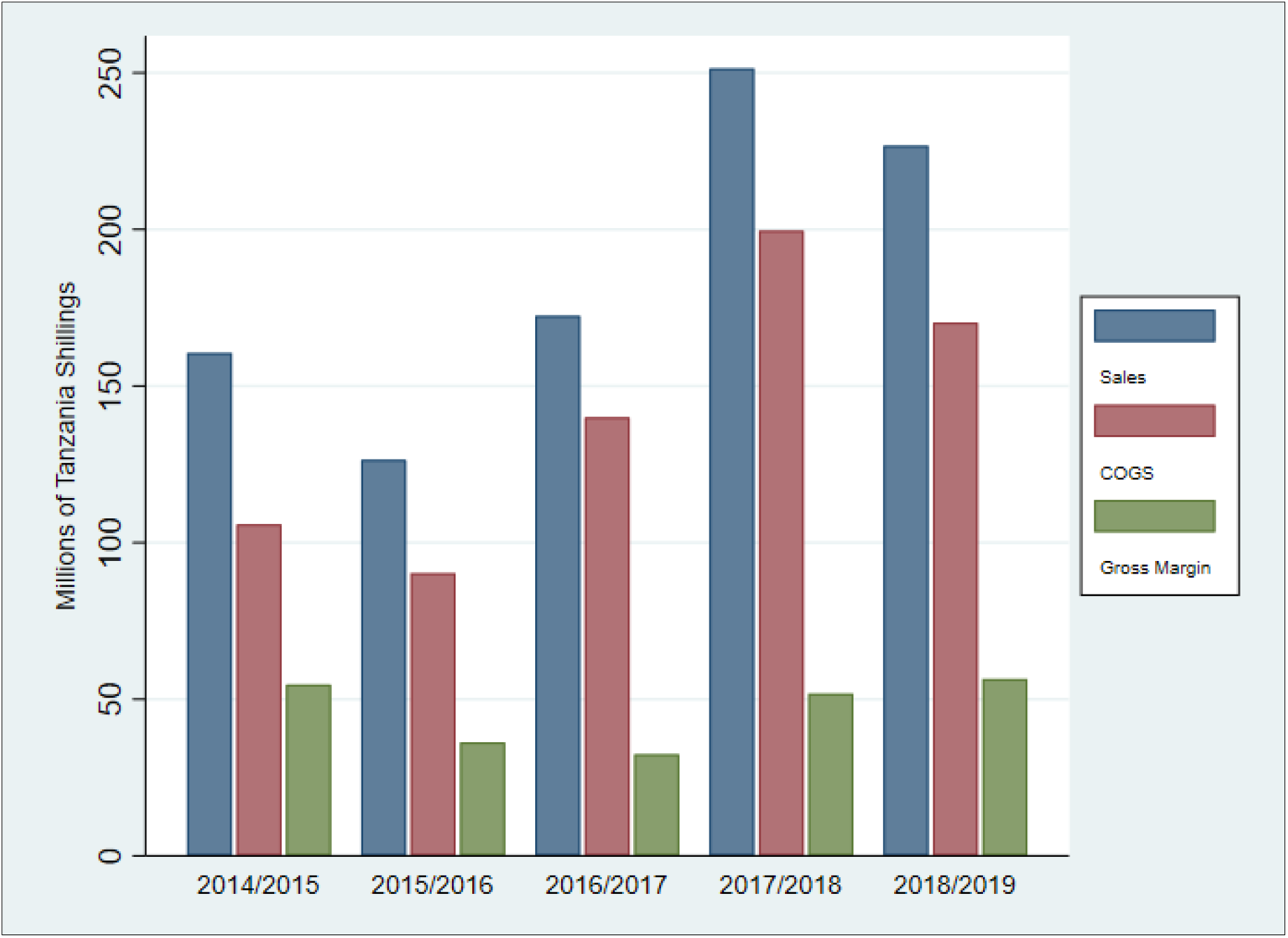
Annual financial performance.

A further examination of the annual income statement shown on Table 4, indicates that other incomes and grants contributed substantially to the financial position of MSD, with an overall increase of 56.4% of the reported surplus between 2014/2015 and 2017/2018 financial period. This period also reported an increase in the total expenses, by 33.2%. According to the granular data from financial reports (not shown), expenses related to employee compensation increased by 38.2%, meanwhile the rental expense reduced by 56.2% over the same period. Fuel costs reduced by 13.0%, while the insurance expenditure increased by 166.3%. Most of the other expenses remained relatively stable over the financial reporting period.

**Table 4:** Income statement.

| Time period | 2018/2019 | 2017/2018 | 2016/2017 | 2015/2016 | 2014/2015 |
| --- | --- | --- | --- | --- | --- |
|  | TZS '000 | TZS '000 | TZS '000 | TZS '000 | TZS '000 |
| **Sales** | 235,352,238 | 251,389,265 | 172,413,432 | 126,440,444 | 160,533,362 |
| **Cost of Goods Sold** | (201,691,890) | (199,605,404) | (140,008,474) | (90,219,538) | (105,835,373) |
| **Gross margin** | 33,660,348 | 51,783,861 | 32,404,958 | 36,220,906 | 54,697,990 |
| **Other incomes and Grants** | 41,792,583 | 36,180,063 | 18,448,061 | 12,325,750 | 5,974,869 |
| **Total expenses** | (54,989,999) | (50,972,685) | (41,596,525) | (43,463,270) | (41,822,737) |
| **Other gains and losses** | (14,938,102) | (27,324,442) | (1,632,470) | 1,454,877 | (12,668,411) |
| **Surplus/(deficit) for the period** | 5,524,830 | 9,666,797 | 7,624,024 | 6,538,263 | 6,181,711 |

Table 5 shows the financial ratios related to the performance of MSD. Between 2014/2015 and 2018/2019, the current ratio averaged 1.5. Meanwhile, over the same period, the cash ratio averaged, 0.3, with the highest being 0.5 reported in the 2016/2017 period. The receivables turnover ratio improved from 1.0 in 2015/2016, to 1.3 in 2018/2019 period, with the highest ratio being 1.6 at 2017/2018. Similarly, the inventory turnover ratio improved from 1.5 in 2015/2016, realizing peak performance of 4.8 in the year 2017/2018. This trend was mirrored by the days inventory outstanding, which reported a reduction from 237 days in the year 2015/2016 to 147 days in the year 2018/2019, with peak performance in 2017/2018 at 76 days.

**Table 5:** Financial Ratios.

| Time Period | 2018/2019 | 2017/2018 | 2016/2017 | 2015/2016 | 2014/2015 |
| --- | --- | --- | --- | --- | --- |
| **Current Ratio** | 1.2 | 1.5 | 1.5 | 1.6 | 1.4 |
| **Cash Ratio** | 0.2 | 0.2 | 0.5 | 0.2 | 0.4 |
| **Receivables Turnover Ratio** | 1.3 | 1.6 | 1.3 | 1.0 |  |
| **Inventory Turnover Ratio** | 2.5 | 4.8 | 3.7 | 1.5 |  |
| **Days Inventory Outstanding** | 147 | 76 | 98 | 237 |  |
| **Net Working Capital Turnover Ratio** | 1.0 | 2.6 | 2.0 | 1.6 | 2.3 |
| **Cash to Cash Cycle** | 309 | 170 | 177 | 253 |  |

The net working capital turnover ratio was stable over the four-year period, averaging 1.9, with the highest performance being 2.7 in 2017/2018. In tandem, the cash to cash cycle was generally high averaging, 227 days over the four-year period. However, from 2015/2016 to 2017/2018, there was a general reduction in the number of days in the cash to cash cycle from 253 days to 170 days respectively. Other results from the related financial reports (shown in Appendix 2), indicated that the working capital increased by 35.1% over the period 2014/2015 and 2017/2018, while the government receivable increased by 50.2%.

### Health Worker Sentiments

Table 6 shows some of the key results from the thematic analysis framework applied in assessing the views expressed by different key informants. Majority of those interviewed acknowledged that the Global Fund support to MSD had been central in improving access to medicines, ramping up efficiency and reliability of its operations. In addition, respondents in the leadership of MSD revealed that the support had benefited the overall financial position of the organization.

**Table 6:** Thematic analysis framework.

| **Codes** | **Emerging Themes** | **Global Themes** |
| --- | --- | --- |
| **Health facilities used to face many stockouts of essential medicines** | Improved management of medicines and commodities | Improved access to medicines and health commodities |
| **Limited storage capacity hindered MSD operations** |  |  |
| **There is an increased portfolio of products available at MSD** |  |  |
| **Modern fleet of vehicles have improved and expanded the reach of MSD** | Reduction in delivery times to health facilities |  |
| **Training of human resources essential for progress to be made** | Satisfied customers across the health system | Improved responsiveness to health system needs |
| **Effective supervision based on key performance metrics is important** | Better communication between zones and health facilities |  |
| **Strategic management office central in tracking progress** | MSD Zones as business units are effective in meeting health facility needs |  |
| **Tracking progress and getting feedback is important for progress** |  |  |
| **Data use is important for effective management decisions** | Improved forecasting and quantification of needs |  |
| **Optimal storage conditions have greatly improved MSD operations** | Reduced inventory loss due to wastage and expiries | Improved efficiency within the healthcare system |
| **MSD can directly procure medicines and commodities from manufacturers** | Cost cutting opportunities due to economies of scale |  |
| **Better visibility of medicines and health commodities across the system** |  |  |
| **There has been expansion of MSD operations to reach every corner of the country** |  |  |
| **There has been a reduction in expenditure on maintenance of old fleet and leasing of warehouses** |  |  |
| **Global Fund has been a strong partner** | Donors and government need to meet their obligations | Sustainability of the gains important |
| **Imperative for timely payment to ensure MSD operations are sustainable** | Improvements in revenue generation capacity towards sustainable operations |  |
| **Need for other donors to support the MSD operations** |  |  |

Majority of the respondents cited the modern warehousing facilities both at the central and zonal levels as good investments that had enabled MSD to attain better visibility and storage of medicines, and other health supplies creating optimal conditions; and thus, enhancing performance. In addition, it was reported by the MSD management personnel that, before the procurement of the new warehouse facilities, MSD used to incur significant rental expenses from private facilities which were not optimal for the storage of priority medicines and other health commodities.

There was also consensus from both the MSD staff and health facilities respondents that the modern fleet at MSD, had significantly improved the delivery of medicines and health commodities to peripheral health facilities. The delivery times were reported to have significantly reduced and health facilities managers were confident to receive their orders on time, and thus reducing the need for overstocking and resultant expiries. The physical conditions of the health commodities at delivery were also reported to have improved, as a result of better storage and transportation conditions. However, it was noted by some MSD staff at the zonal level, that the available fleet was not optimally utilized because of the limited number of drivers that had been employed.

Most of those interviewed also reported that training of human resources had been augmented through the Global Fund support and this has been a critical pillar in improving the performance of MSD. Majority of respondents among the MSD staff reported that they were more aware of their roles and responsibility since receiving the necessary training and sensitization. This awareness was further reinforced by the regular monitoring and evaluation that had been institutionalized through the SMO. MSD senior management respondents pointed out the board level reforms that had been instituted through the Global

Fund and USAID support as being critical to the overall strategic leadership and effective management of the organization.

Many of the respondents agreed that quality data were essential for the smooth functioning of procurement and supply chain systems. Therefore, they cited the investments in the information management systems implemented through the Global Fund support as having played a key role in improving performance at the organization. The MSD management revealed that through better collection and use of high-quality data, decision makers at various levels were empowered to effectively quantify and forecast needs as well as have better visibility of distribution systems.

At the zonal and facility levels, the investment in information management systems had reportedly allowed for better visibility of products and seamless ordering of medicines and health commodities, from higher levels or redistribution whenever necessary. Many respondents concurred that this situation had led to reduced expiries and wastage of essential medicines and other health commodities, which previously were common occurrence.

According to the MSD management, its financial position had been significantly strengthened by the fact that Global Fund regularly paid 6% of the cost of vertical program’s products it channeled through MSD’s distribution network. This formed a substantial revenue stream that is critical in ensuring the financial sustainability of MSD. However, it was also reported that other development partners that were channeling health commodities through the MSD distribution network were not making similar commitments, a situation that put the organization’s sustainability at risk. Similarly, the government had accumulated significant receivables to MSD, if not fully settled on time, this would significantly disrupt the smooth operations at the organization.

## Discussion

Our study substantially contributes to the literature on strengthening health systems in low- and middle-income countries, with specific emphasis on procurement and supply chain systems. This is an important subject of inquiry considering that many countries are still grappling with health service delivery gaps, that could be directly or indirectly attributed to dysfunctional procurement and supply chain systems [15-17, 19].

In interpreting the results presented in this article, we are cognizant that we have based our findings on limited observations, and over a brief period. We are also aware that our study could be subject to confounding relationships which could complicate our attribution of impact to specific interventions. However, we have made efforts to mitigate for this, by employing a mixed methods approach, using both quantitative and qualitative research methods, and triangulating multiple data sources into a consistent analytical framework.

There is consistent evidence that the Global Fund supported reform resulted in positive improvements in the overall performance at MSD. Availability of essential medicines and other health commodities increased, indicating improved capacity to meet demands emanating from the health facilities. This could be attributed to several factors related to the Global Fund support; notable ones being, better quantification and forecasting capabilities (due to availability of data and training); improved warehousing capacity to hold a wide portfolio of products and direct delivery to facilities through a modern fleet. Other studies point to the importance of such interventions [19, 25, 26].

Further, the newly formed SMO, was impactful in driving institutional reforms such as strengthening the board oversight capacity, human resource training and supervision. More specifically, SMO was central in the institutionalization of the culture of performance assessment, driving change in governance and advocating for the adoption of information system improvements and prudent financial management. All these measures were deemed contributory to the observed improvements reported at MSD [9].

Further, through Global Fund support, MSD had increased capacity procure and store an expanded portfolio of products, necessitating the expansion of tracer items routinely tracked from 135 to 312. In addition to the operational improvements, MSD became in better financial position to negotiate preferential procurement terms directly from manufacturers, attracting better prices and a wider range of products to meet public health needs. However, despite these enhanced capabilities, it was concerning that the order fill rates from both the headquarters to the zones and the zones to health facilities, had not shown significant improvement. Some of the reasons for this was because old products that had optimal availability at facilities, were removed from the tracer list and new ones introduced. Further, direct sourcing from manufacturers initially presented a challenge for many new items did not have an active framework agreement. This has now been addressed and improvements are expected.

Through an expanded ILS system, personnel at the central and zonal levels have improved visibility of the current needs at respective health facilities and are empowered to rationalize the distribution of products to meet the needs on the ground. With improved operational performance, the financial position of MSD showed a growth trajectory. The increased revenues could be attributed to several factors including, a steady revenue stream from the Global Fund service fees and an improved warehousing infrastructure which allowed MSD to procure and store an extended range of products for sale. In addition, the distribution network to reach more health facilities had been strengthened through the modern fleet, and this translated into more sales. The Government of Tanzania had also increased its allocation for procurement of health products, as a result of the advocacy efforts by various stakeholders and its commitments to meet global health goals [3,7,9].

Increased income surplus at MSD was a positive indication towards achieving financial sustainability. In addition to increasing sales, the relative reduction in key expense items like rent for warehousing and maintenance for aging equipment and vehicles were critical, and these had been realized through Global Fund support. The current ratio which underscores the liquidity position of MSD to meet its short-term financial obligations was generally favorable, with a gradual improvement over time. This could be attributed to the measures implemented at MSD such as implementing a cash and carry policy and efforts to collect overdue receivables from key accounts, with a varying level of success [9].

Despite the improvements in the successive current ratios, the more stringent liquidity measure, cash ratio was generally, low across the years. A cash ratio of between 0.5 and 1 is considered good, but in the case of MSD, the average was, 0.31, indicating that MSD might not be able to sufficiently meet its short-term financial obligations, using cash and cash equivalents at its disposal. This was a significant risk to MSD, given the nature of its operations, where rapid procurement might be needed to respond to emergencies or steady commitments, were needed from suppliers and manufacturers, to ensure minimal disruptions in service delivery.

Meanwhile, the receivable turnover ratio, was low despite the marginal increase over the years; indicating that MSD was not quickly converting the credit sales into cash to support its operations. In fact, the increasing government receivable, which increased by more than 50% between 2014/2015 and 2017/2018 was a major risk to MSD’s financial solvency and overall sustainability. Another key pointer to this trend was the long cash to cash cycle, which despite an improving trend, was still significant. On average it took 227 days to convert the inventory held by MSD into cash. It was very clear that a significant portion of this cash value was locked within the ballooning government receivable.

The improvements in inventory turnover ratios over the years, showed better capacity to effectively manage inventory, and hence reduction in potential losses due to wastage, obsolesce and expiries. Better warehousing, transportation, enhanced product visibility, responsive and empowered zonal management teams, among others, were all critical to these observed progress. But still, it must be clarified that, despite the improvements in inventory management, more still needed to be done. On average, it took MSD 140 days, to sell its inventory, which led to significant holding costs, eroding the bottom-line.

Overall, the role of the donor community and the government is still central to the future sustainability of MSD. Therefore, in order to strengthen MSD’s financial position, development partners and other stakeholders including government, which distribute medicines and health commodities through the MSD channels should be cognizant of the cost implications associated with these activities. All stakeholders must undertake to pay their fair share of the related costs in a timely manner in order to avoid interruptions to MSD operations.

Ultimately, for MSD to effectively meet its obligations and deliver on its mandate, it must be led by an accountable team focused on continually improving its performance. The existing governance framework comprising of an independent board staffed by high caliber professionals and an experienced management team at the helm of MSD provides an opportunity for further transformation. This will sustain the momentum generated by the reform process and enable MSD to become an exemplar self-sustaining organization for others to emulate.

## Conclusions

In summary, there is clear evidence that the Global Fund supported reforms at MSD yielded the desired impact and are on track to achieving the performance and sustainability objectives. However, there are still many risks that could derail this trajectory and should be anticipated and adequately addressed.

The importance of an effective procurement and supply chain system to support health service delivery cannot be overstated. It is a critical ingredient to make progress towards universal health coverage [1,2, 15-19]. The Tanzania MSD example demonstrates that in order to build effective procurement and supply chain systems, sustained and multifaceted support is necessary as opposed to piecemeal and isolated undertakings. Effective supportive measures need to involve all the key health system stakeholders, including those involved in the financing, planning and implementation of health programs [1,2].

## Data Availability

All data used in this manuscript are available upon request and with permission from Tanzania MSD

## Authors’ contributions

TA, LM, PG and SP conceptualized and designed the study. TA, RS and ED carried out data collection and analysis. TA and PG drafted the manuscript and LM, SP, RS, NM, TP, SM, AL and LB did the critical revisions of the manuscript. All authors read and approved the final manuscript.

### Acknowledgements

We would like to thank the participants from the different organizations that were interviewed and provided feedback during the study. We are also grateful to the management of the various organizations, particularly MSD that allowed their staff to participate and provided premises and other forms of facilitation during the study. More specifically, we appreciate the facilitation support from Pascal Pastory, Salome Mallamia, and Bwekwaso Tabura, all from MSD.

## Data sharing

All raw material from the key informant interviews and secondary data analysis may be made available by the authors upon request; and with written permission from MSD and MoHCDGEC.

## Dissemination declaration

The research findings shall be disseminated to the Tanzania healthcare system stakeholders, including MSD staff

---

**Appendix 1:**
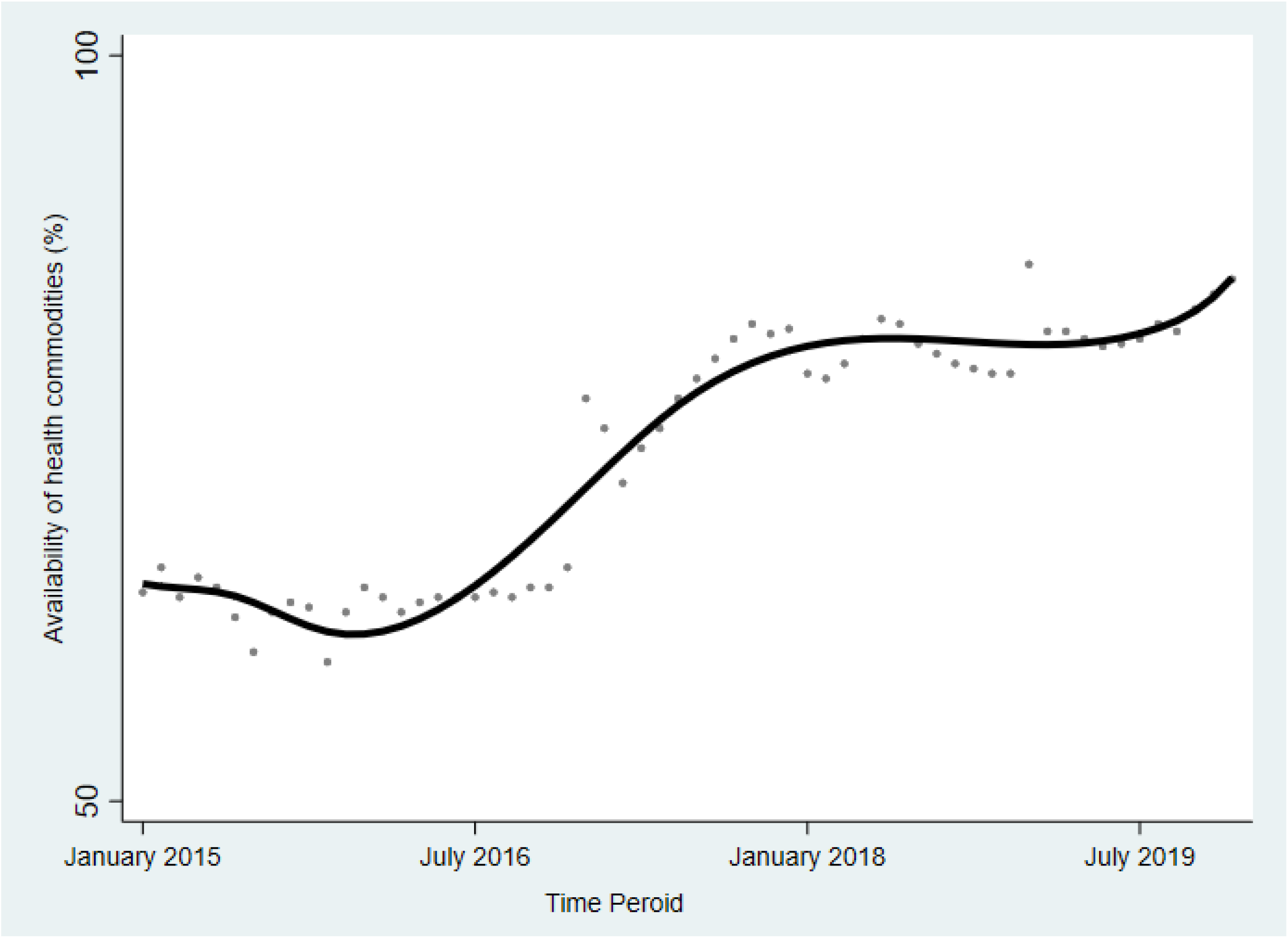
Financial Statement.

**Appendix 2:**
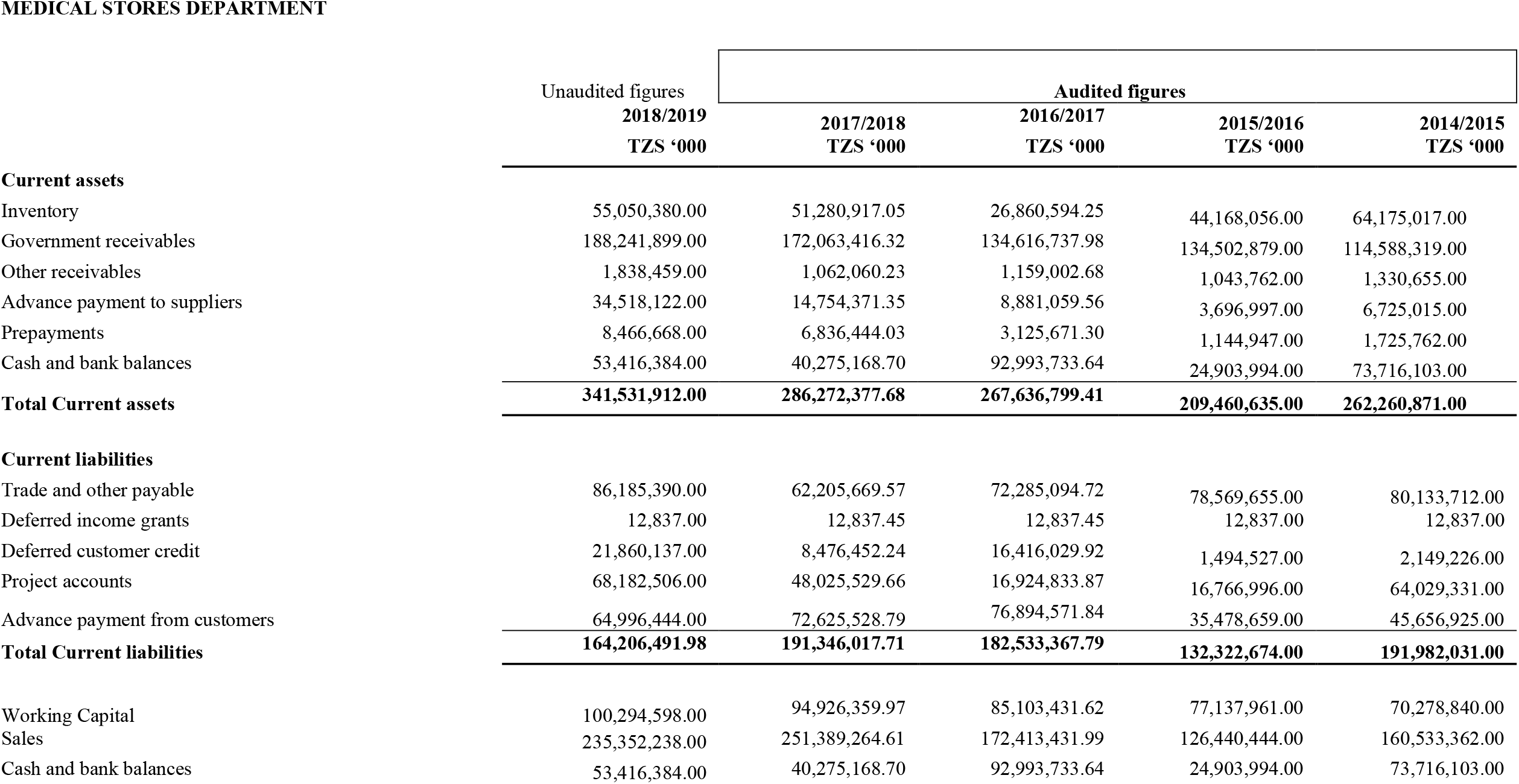
Flexible cubic spline model: availability of health commodities over time.

